# Comprehensive analysis of SARS-CoV-2 antibody dynamics in New Zealand

**DOI:** 10.1101/2020.12.10.20246751

**Authors:** Alana L. Whitcombe, Reuben McGregor, Alyson Craigie, Alex James, Richard Charlewood, Natalie Lorenz, James M.J. Dickson, Campbell R. Sheen, Barbara Koch, Shivani Fox-Lewis, Gary McAuliffe, Sally A. Roberts, Susan C. Morpeth, Susan Taylor, Rachel H. Webb, Susan Jack, Arlo Upton, James Ussher, Nicole J. Moreland

**Affiliations:** Faculty of Medical and Health Sciences, University of Auckland, New Zealand; Maurice Wilkins Centre, University of Auckland, New Zealand; Southern Community Laboratories, Dunedin, New Zealand; Te Punaha Matatini and School of Mathematics and Statistics, University of Canterbury, Christchurch, New Zealand; New Zealand Blood Service, Auckland, New Zealand; School of Biological Sciences, University of Auckland, New Zealand; Protein Science and Engineering, Callaghan Innovation, Christchurch, New Zealand; LabPLUS, Auckland City Hospital, Auckland, New Zealand; Middlemore Hospital, Auckland, New Zealand; Starship Children’s Hospital and Kidz First Children’s Hospital, Auckland, New Zealand; Public Health South, Southern District Health Board, Dunedin, New Zealand; Department of Microbiology and Immunology, University of Otago, Dunedin, New Zealand

**Author notes:** **Corresponding Author:** Dr Nicole J. Moreland, Department of Molecular Medicine and Pathology, Faculty of Medical and Health Sciences, University of Auckland, Private Bag 92019, Auckland, New Zealand.

**Keywords:** COVID-19, SARS-CoV-2, immunokinetics, neutralising antibodies, Spike protein

## Abstract

**Objectives:** Circulating antibodies are important markers of previous infection and immunity. Questions remain with respect to the durability and functionality of SARS-CoV-2 antibodies. This study explored antibody responses in recovered COVID-19 patients in a setting where the probability of re-exposure is effectively nil, owing to New Zealand’s successful elimination strategy.

**Methods:** A triplex bead-based assay that detects antibody isotype (IgG, IgM and IgA) and subclass (IgG1, IgG2, IgG3, IgG4) responses against Nucleocapsid (N) protein, Receptor Binding Domain (RBD) and Spike (S) protein of SARS-CoV-2 was developed. After establishing baseline levels with pre-pandemic control sera (n=113), samples from PCR-confirmed COVID-19 patients with mild-moderate disease (n=189) collected up to eight months post-infection were examined. The relationship between antigen-specific antibodies and neutralising antibodies (NAbs) was explored with a surrogate neutralisation assay that quantifies inhibition of the RBD/hACE-2 interaction.

**Results:** While most individuals had broad isotype and subclass responses to each antigen shortly after infection, only RBD and S protein IgG, as well as NAbs, were stable over the study period, with 99%, 96% and 90% of samples, respectively, having responses over baseline 4-8 months post-infection. Anti-RBD antibodies were strongly correlated with NAbs at all time points (Pearson’s r ≥ 0.87) and feasibility of using finger prick sampling to accurately measure anti-RBD IgG was demonstrated.

**Conclusion:** Antibodies to SARS-CoV-2 persist for up to eight months following mild to moderate infection. This robust response can be attributed to the initial exposure without immune boosting given the lack of community transmission in our setting.

## Introduction

It is now well established that antibody responses against Severe Acute Respiratory Syndrome Coronavirus 2 (SARS-CoV-2) are activated promptly after infection.^1^ The virus, and causative agent of the Coronavirus Disease 2019 (COVID-19) pandemic, contains four structural proteins, the most immunodominant being the Nucleocapsid (N) protein, Spike (S) protein and the receptor binding domain (RBD) of the S protein. Measuring antibody responses to these antigens using serological assays has been critical for determining previous viral exposure in individuals, studying community transmission, and conducting population sero-surveys.^1^ However, much is still to be learnt about the long-term duration and protective capacity of these responses.

Antibody responses comprise different isotypes and subclasses, each associated with unique immune functions and dynamics over time.^2^ Following SARS-CoV-2 infection there is an almost concurrent rise in Immunoglobulin M (IgM), IgA and IgG, with IgM then beginning to decline approximately three weeks after symptom onset.^3–6^ There have been conflicting reports with respect to IgG duration, ranging from a relatively short three months^7^, to six months or longer^8–10^, in part due to SARS-CoV-2 antibody dynamics being highly antigen-dependant. Anti-N antibodies are now known to wane faster than anti-RBD and anti-S, and may be more suited as a marker of recent COVID-19 infection, particularly since the N protein is associated with RNA packaging and anti-N antibodies are non-neutralising.^8,11^ The RBD of the Spike protein on the other hand binds to the Angiotensin-Converting Enzyme-2 (hACE2) on human host cells to facilitate viral entry and infection. Anti-S and anti-RBD antibodies can block this interaction leading to viral neutralisation, and as such, are better markers of functional immune responses.^12^ Of note, the presence of neutralising antibodies (NAbs) protected a small number of individuals from re-infection during a SARS-CoV-2 outbreak on a fishing vessel^13^ and anti-S protein IgG was associated with reduced re-infection in a recent study of health care workers in the United Kingdom.^14^ This suggests NAbs and S protein antibodies are associated with protective immunity and may, in turn, underpin a correlate of protection for COVID-19 vaccine development. Levels of NAbs have proven relatively stable in recent reports^9,10,15^ but further investigation is needed, particularly in non-severe cases of COVID-19.

Bead-based serological assays capable of simultaneously measuring antibodies to N and S proteins, together with RBD, enable a comprehensive view of the SARS-CoV-2 antibody response.^16–18^ In this study, a triplex Luminex-based assay that detects isotype and subclass responses to the major SARS-CoV-2 antigens was developed and utilised to interrogate the composition and duration of virus-specific antibodies up to eight months post-infection in COVID-19 cases in New Zealand. In parallel, levels of NAbs were measured using a surrogate Virus Neutralization Test (sVNT) previously shown to strongly correlate with neutralisation measured using live SARS-CoV-2 virus, and better suited to high throughput analyses due to improved speed and reproducibility.^12,19^ By exploring the relationship between NAbs and antigen-specific antibody features, this study adds to growing knowledge of the SARS-CoV-2 humoral immune response from a setting where the probability of re-exposure is effectively nil, owing to New Zealand’s successful elimination strategy.^20,21^

## Results

A triplex bead-based immunoassay was developed, with N protein, trimeric S protein and RBD coupled to spectrally unique beads. Compatibilty of the beads in a multiplex format was confirmed, as was comparability with previously described ELISA for S protein and RBD^***22***^, and N protein titres determined using the Abbott Architect SARS-CoV-2 IgG assay^***23***^, with highly significant correlations for all three antigens (Supplementary Figure 1). The level of SARS-CoV-2 antigen specific isotypes (IgG, IgM, IgA) and subclasses (IgG1, 2, 3 and 4) were determined in a cohort of 112 PCR-confirmed COVID-19 participants, 50 of whom had multiple time points (n=189 samples), and the majority of whom had mild disease (Table 1). The days post-infection ranged up to 246 days, enabling the kinetics of 21 different antibody features (3 SARS-CoV-2 antigens and 7 secondary detectors) to be temporally investigated. The cut-off for positivity for each antibody type was determined using a previously described panel of pre-pandemic control samples (n=113) that includes donors with respiratory viruses and bacterial pneumonia that have symptom overlap with COVID-19.^***22***^

**Table 1.** Demographics of COVID-19 study participants

| Demographic characteristic | COVID-19 patients, grouped by days post symptom onset |  |  |  |
| --- | --- | --- | --- | --- |
|  | <7 days | 7-62 days | 63-124 days | 125-250 days |
| Total samples, *n* | 3 | 27 | 79 | 80 |
| Age (year) |  |  |  |  |
| Median | 61 | 41 | 51 | 50 |
| Range | 29 - 64 | 23 - 86 | 17 - 81 | 17 - 81 |
| Gender, *n* (M/F) | 0/3 | 14/13 | 33/46 | 31/49 |
| **Clinical severity** |  |  |  |  |
| Admitted to hospital with moderately severe/severe disease, *n* (%) | 3 (100) | 18 (66.7) | 4 (5.1) | 1 (1.3) |

**Figure 1.**
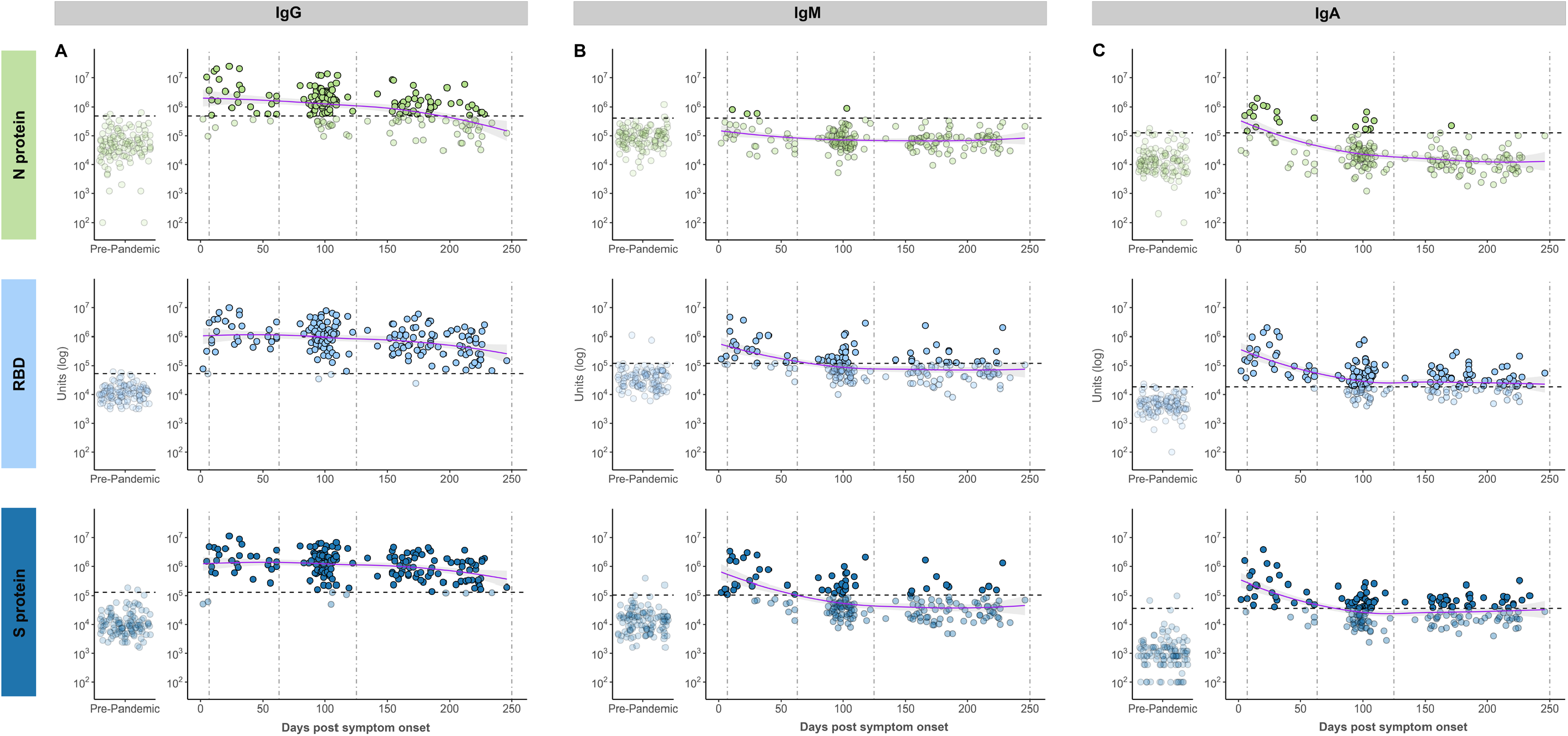
Isotype responses for IgG (A), IgM (B) and IgA (C), against each antigen (Nucleocapsid (N) protein in light green, receptor binding domain (RBD) in light blue, Spike (S) protein in dark blue). Left panel shows responses in pre-pandemic controls (n=113), right panel shows responses in PCR-confirmed COVID-19 patient samples (n=189) over time. Dashed vertical lines represent the three time groups (7-62 days, 63-124 days, 125-250 days). Dashed horizontal lines represent baseline cut-off’s calculated from the pre-pandemic control group and samples below these cut-offs are faded. LOESS regression (blue line) was used to visualise general trends in antibody levels, and standard error of regression indicated by grey shaded area.

Isotype responses for each antigen were plotted against time (Figure 1). Participants were grouped by days/months post-infection (7-62 days (0-2 months, n=27), 63-124 days (2-4 months, n=79) and 125-250 days (4-8 months, n=80)) and the rate of positivity compared (Supplementary Table 1). With the exception of anti-N protein IgM, all antigen-specific isotypes were elevated above baseline levels in the acute stage through to early convalescence (7-62 days post symptom onset). Indeed, all samples (27/27, 100%) were positive for RBD and S protein IgG indicating universal seroconversion following infection in these participants. After two months, IgM and IgA antibody levels against all three antigens trended down to baseline levels. In contrast, IgG antibody levels remained elevated against RBD and S protein with 99% (79/80) and 96% (77/80) of samples above baseline in late convalescence (≥125 days), respectively. Consistent with recent literature^8,11^, anti-N protein IgG waned faster, with only 54% (43/80) of patients having detectable levels above baseline in late convalescence.

To explore the contribution of IgG subclasses to total IgG (IgG_tot_), the proportion of samples with antibody subclass levels above baseline in the three time groups were compared (Figure 2, Supplementary Table 1). The four IgG subclasses were elevated against each antigen in the majority of samples collected 7-63 days post-symptom onset. IgG3 responses were particularly strong in this earliest time group, with at least 96% of patients having a response above baseline against both RBD and S protein (Supplementary Table 1). In samples collected 63-124 days post symptom onset, IgG1 and IgG3 responses dominated for all three antigens, with IgG2 uncommonly, and IgG4 rarely, above baseline. In the 125-250 day group IgG1 antibodies against RBD and S protein persisted, with 64% (51/80) and 59% (47/80) of participants having antibodies above baseline, respectively, as did IgG3 against S protein with 60% (48/80) above baseline. As with IgG_tot,_ the anti-N subclass responses waned faster than RBD and S protein with <30% participants having detectable levels above baseline of any subclass in the 125-250 day group.

**Figure 2.**
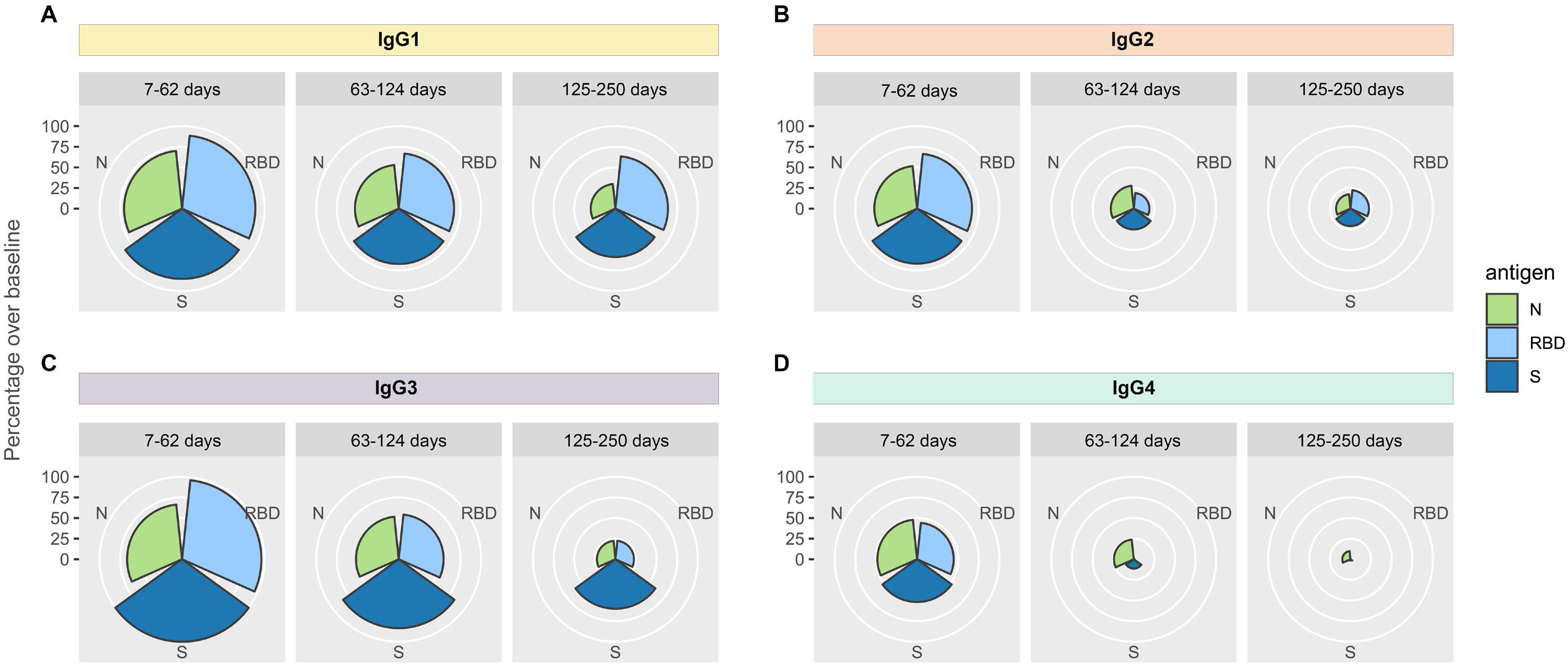
Rose plots showing the percentage of PCR-confirmed COVID-19 patient samples, that were above baseline for IgG1 (A), IgG2 (B), IgG3 (C) and IgG4 (D). Samples are stratified into three time groups (7-62 days, 63-124 days, 125-250 days). Nucleocapsid (N) protein in light green, receptor binding domain (RBD) in light blue and Spike (S) protein in dark blue.

NAbs were measured using a surrogate assay that quantifies inhibition of the RBD/ACE-2 interaction^12^ and found to be relatively stable over the eight month study period (Figure 3). While there was a modest downward trend, 89% (70/79) in the 63-124 day group and 90% (72/80) in the late convalescent group (>125 day) had detectable NAbs above the cut-off. All bar one of the late convalescent samples (79/80) were from non-hospitalised individuals suggesting mild disease can induce long-lived NAbs. Correlation analysis found IgG_tot_-RBD most significantly correlated with NAbs in the early, mid- and late-convalescent groups (Pearson’s r ≥ 0.87 across all time groups) while IgA-RBD was moderately associated, and IgM-RBD not at all (Figure 3B). To further explore NAb persistence, a comparative analysis was performed on participants for which two or more samples were collected across the study period (Supplementary Figure 2). There was no significant decrease in NAbs between the first and second time point, but a significant decline at the third time point (median 219 days post-infection, *P* < 0.05). Individual heterogeneity was evident in terms of both the level of NAbs, and their waning (or otherwise) over time. Indeed, approximately 1/3 of individuals demonstrated increasing levels, consistent with affinity maturation as recently proposed.^24^

**Figure 3.**
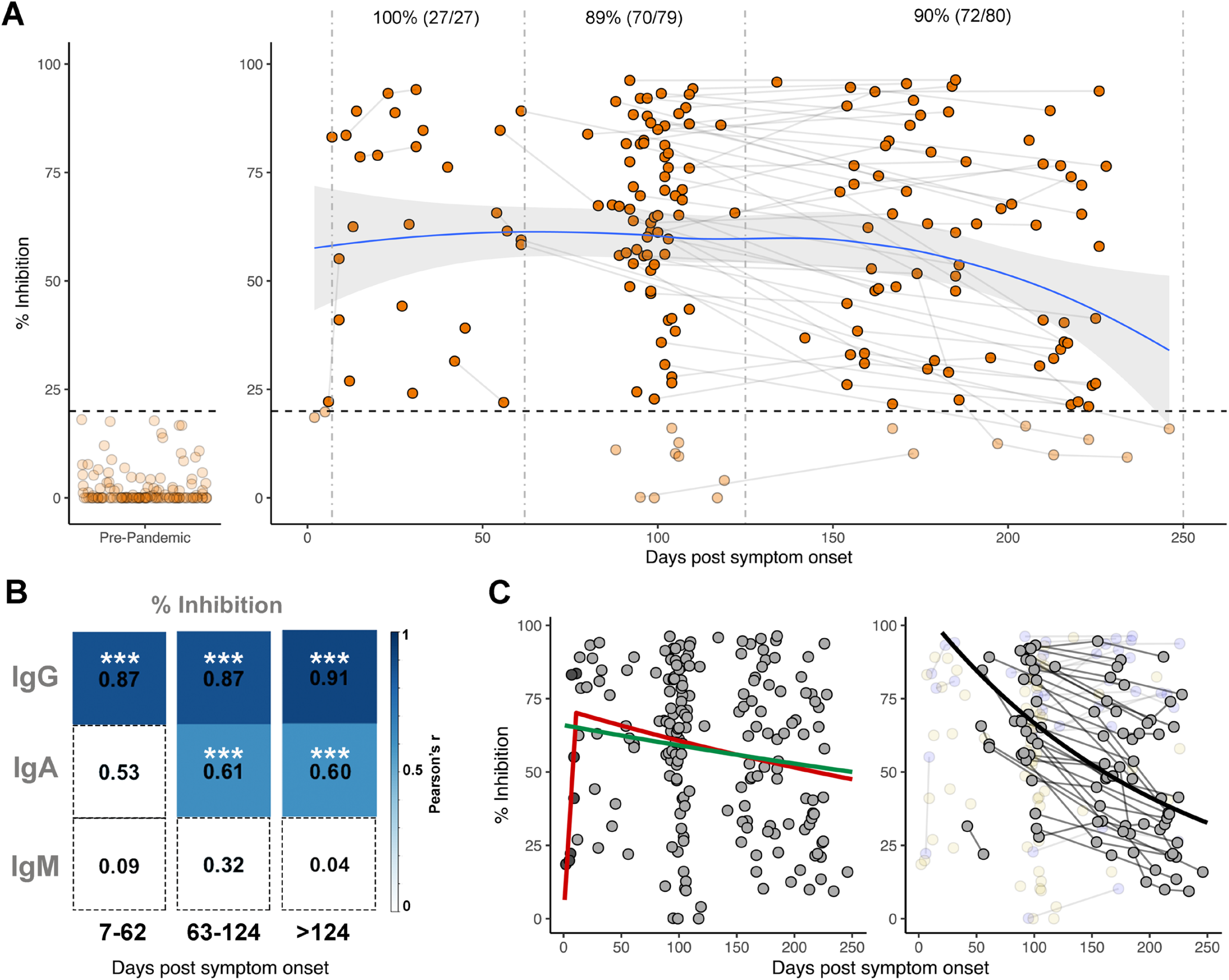
Longitudinal neutralising antibodies. (A) Neutralising capacity (in % inhibition) of samples from days since symptom onset. Left panel shows responses in pre-pandemic controls (n=113), right panel shows responses in PCR-confirmed COVID-19 patient samples (n=189) over time. Pre-pandemic samples were all below the 20% cut-off indicated by dashed horizontal line. Paired samples are joined by grey lines. Dashed vertical lines represent the three time groups (7-62 days, 63-124 days, 125-250 days). LOESS regression (blue line) was used to visualise general trend in neutralising antibody levels, and standard error of regression indicated by grey shaded area. Percent and number of samples from PCR-confirmed cases above 20% inhibition cut-off is indicated above the respective time points. (B) Pearson correlation between % inhibition vs RBD IgG, IgA and IgM across three time groups. Non-significant correlations are coloured white. Significant correlations are coloured in blue relative to their Pearson’s r value. *P*-values are corrected for multiple comparisons using Bonferroni method, *** = *P* <0.001. (C). Visualisation of three modelling methods applied to predict half-life of neutralising antibodies. Left panel shows the “Exponential decay” (green line) and “growth and decay” (red line) models which use all samples to model decay. In the “growth and decay” model, samples up to 11 days (dark grey points) are used to model the growth phase, and samples over 11 days (light grey) are used to model exponential decay. Right panel shows the “individual decay” model (black line) in which only samples with multiple paired measurements who were in the decay stage were utilised (grey points). Samples with only one measurement (yellow faded points) or those showing increased inhibition over time (blue faded points) were excluded.

To estimate the rate of decay for NAbs a series of models were applied (see methods), and the expected time for NAbs to halve (half-life (t_1/2_)) was calculated (Figure 3C, Supplementary Table 2). Exponential decay assumes NAbs decline immediately after infection, and estimated the longest t_1/2_ of 625 days and the largest margin of error (95% CI, 319-13465 days). A growth and decay model that assumes NAbs increase initially and then decline, estimated a shorter t_1/2_ of 425 days (95% CI, 253-1316). Using only data from the 50 participants for which two or more samples were available, excluding those with increasing NAbs (i.e. still in the growth phase), and modelling individual decay, estimated a much shorter t_1/2_ of 146 days (95% CI, 100-199).

Lastly, since IgG_tot_ persist, and the bead-based assay format is compatible with small quantities of sera (2μL), the feasibility of using dried blood samples to measure IgG_tot_ was explored. Dried blood samples could expand the acceptability of serological assays and enable testing in settings where it maybe logistically challenging to collect, process and store venous blood.^***25***^ Simulated dried blood eluents collected using Mitra samplers were compared to matched sera in a subset of patients (n=19). IgG_tot_ measurements from Mitra eluents correlated strongly with serum samples, against all three antigens (r^2^ = 0.9928, 0.9964 and 0.9931 for N, RBD and Spike, respectively) (Figure 4).

**Figure 4.**
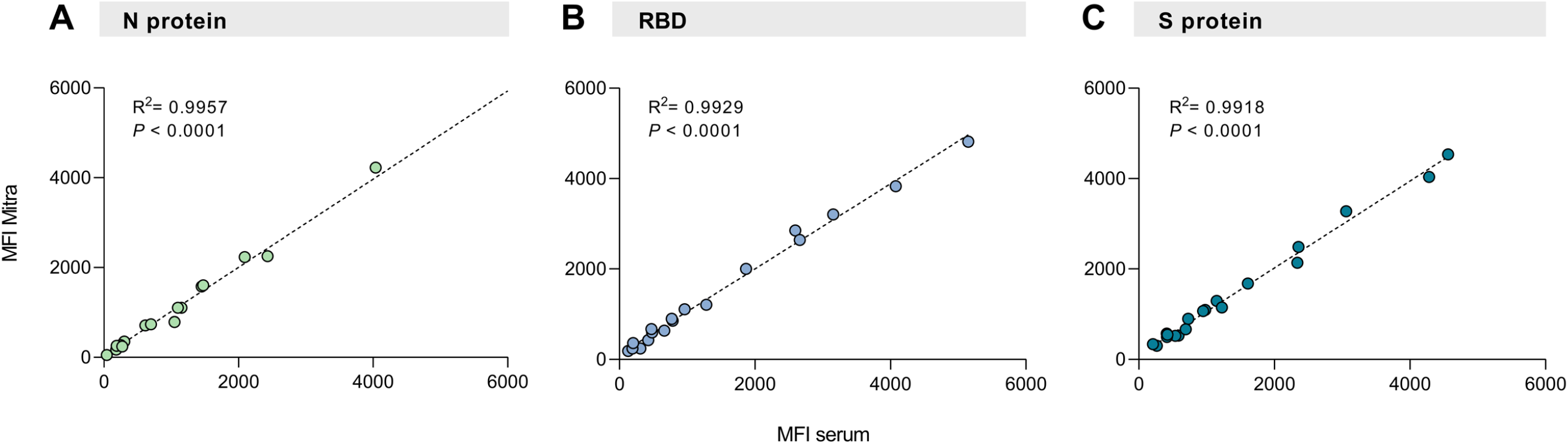
Scatterplots showing the correlation between IgG measured from Mitra dried blood eluents and matched serum for Nucleocapsid (N) protein (A), receptor binding domain (RBD) (B) and Spike (S) protein (C) (n=19) in median fluorescence intensity (MFI). The dashed lines represent the linear regression equation. R squared and P-values are shown.

## Discussion

The duration of antibody responses following SARS-CoV-2 infection is, in essence, being studied in real-time globally. Initial studies that purported early antibody waning were based on the N protein, now known to induce antibodies with a relatively short half-life^8^, or linear extrapolations of S protein data collected 2-3 months post-infection.^7^ More recent analyses (5-8 months post-infection) show that S protein, RBD and NAbs are likely more stable than originally predicted.^6,8,10,15,18,26^ Here, further evidence is provided of the durability of the SARS-CoV-2 humoral immune response over an eight month period. Importantly, this study was performed in a setting where there is next to no likelihood of immune boosting by re-exposure, given the successful public health response to COVID-19 in New Zealand.^20,21^ That 99% of sera had anti-RBD IgG and 96% had anti-S protein IgG above baseline levels 4-8 months after infection in a setting with no circulating SARS-CoV-2 is notable, particularly given the majority of participants had non-severe infections.

Utilising a multiplex assay enabled not only IgG, but other isotypes and subclass responses to be investigated. As expected, IgM had a shorter duration than IgG, with >75% of samples returned to baseline in late convalescence for all three antigens. Of the IgG subclasses, RBD and S protein specific IgG1 and IgG3 were dominant, in keeping with the association of these subclasses with convalescence and survival in COVID-19 systems serology studies.^16,17^ While it is not possible to link subclass responses with disease outcomes in this study since all participants recovered, IgG1 and IgG3 are the most potent subclasses in terms of immune cell engagement and effector functions.^2^

There are still many unknowns with respect to correlates of protection for SARS-CoV-2 infection including the contribution of cross-reactive memory T-cells alongside B-cell driven antibody responses.^27^ For other viral infections correlates are based on specific levels of antibodies, with these well-defined titres facilitating vaccine development and immunisation strategies.^15^ As such there is a pressing need to understand antibody persistence and time to seroreversion following SARS-CoV-2 infection. Using three different models the estimated t_1/2_ of 5-20 months for NAbs in this study is somewhat longer than 2-4 months calculated in other cohorts^18,26^, though all estimates have wide confidence intervals due to the inherent limitations of modelling responses with marked individual heterogeneity. Nevertheless, all models place the NAb half-life at <2 years post-infection. Although NAbs are not yet a proven correlate of protection, and the impact of slow waning on susceptibility to re-infection is as yet unknown, NAbs and anti-S protein IgG have been associated with protection in recent outbreaks^13,14^, and in non-human primate studies^28^ suggesting a role for functional antibodies in SARS-CoV-2 immunity. Ultimately, large-scale vaccine trials will enable accurate determination of a correlate, and suggestions that two-dose Spike-based vaccines may induce more potent NAbs than natural infection^29^ are likely to result in responses of increased durability and longer half-life than estimated to date.

Overall, this study provides a comprehensive view of SARS-CoV-2 antibodies over eight months. The strong correlation between anti-RBD IgG and NAbs, combined with the demonstration in this study and by others^4,25,30^ that anti-RBD can reliably be measured from dried blood fingerpick samples, provides feasibility for future SARS-CoV-2 immunokinetic studies that incorporate RBD-IgG based assays. The importance of conducting such studies at scale during vaccine roll-out is particularly relevant in settings like New Zealand, where there is potential to gain novel insight on vaccine responses given the lack of circulating SARS-CoV-2 in the community.

### Methods Study samples

Plasma and serum samples were collected from multiple sources with appropriate ethical board approval. Samples collected before the pandemic were used as negative controls (“pre-pandemic,” n=113), details of which have been described previously.^22^ Samples from PCR-confirmed COVID-19 individuals were obtained from hospital in-patients in Auckland (n=18) (ethics HDEC 20NTB76) and convalescent participants in the Southern Region of New Zealand (ethics HDEC 20NTB101) as previously described.^22,23^ Samples were also obtained from donors with a prior COVID-19 diagnosis, collected at the New Zealand Blood service as part of a Medsafe approved process for convalescent plasma preparation. The final PCR-confirmed cohort in this study comprised 189 samples from 112 participants, 50 of whom had samples collected at multiple time points (Table 1). Participant samples were grouped by days/months post symptom onset (infection) based on timeframes in which the samplers were obtained (acute and early convalescence, 7--62 days (0-2 months, n=27); mid-convalescence, 63-124 days (2-4 months, n=79); late convalescence, 125-250 days (4-8 months, n=80)).

### Antigen coupling to beads

SARS-CoV-2 RBD and the trimeric S protein antigens were recombinantly expressed and purified from HEK293T or HEK293F cells as previously described.^22^ Recombinant nucleocapsid (N) protein expressed in Baculovirus Insect cells was obtained commercially (SinoBiological). Each of the three antigens were coupled to MagPlex® magnetic microspheres (beads) by carbodiimide chemistry using the xMAP® Antibody Coupling Kit (Luminex Corporation), according to the kit instructions. In brief, beads were washed with activation buffer and incubated for 20 min with EDC (1-Ethyl-3-[3-dimethylaminopropyl] carbodiimide hydrochloride) and Sulfo-NHS (N-hydroxysulfosuccinimide). Antigens were coupled at a concentration of 4.5 μg per 1 × 10^6^ beads in a 2 hour incubation at room temperature. Antigen-coupled beads were washed, enumerated using a haemocytometer, and stored at 4°C protected from light until further use.

### Multiplex assay protocol

All serum and plasma samples were heat-inactivated at 56°C for 30 min prior to use. Samples were diluted 1:100 (IgG2, IgG3, and IgG4), 1:400 (IgG1, IgA, IgM) or 1:800 (IgG) in assay buffer (AB) consisting of PBS + 1% IgG-free bovine serum albumin (BSA, MP Biomedicals). Diluted samples (30 μl) were added to wells of a 96-well U-bottom plate (Greiner) and 30 μl of bead solution consisting of N, RBD and S-coupled beads mixed in equal parts were added at a concentration of 40 beads/μl/antigen. Plates were sealed and incubated at room temperature for 35 min at 800rpm, followed by two wash steps with wash buffer (AB + 0.05% Tween) using a handheld magnet (Luminex Corporation). To detect the different antibody isotypes and subclasses, phycoerythrin (PE)-labelled donkey anti-human detection antibodies (IgG1, 2, 3 and 4, Southern Biotech; IgG/A/M, Jackson Immunoresearch) were diluted 1:75 and 1:80 in AB, respectively, then added to wells and incubated at room temperature for 35 min at 800rpm. Following another two wash steps, beads were re-suspended in 100 μl Drive Fluid (Luminex Corporation), and analysed on a MagPix® machine (Luminex Corporation).

Positive COVID-19 sera (n=10) from a commercially available reference panel (AccuSet™ SARS-CoV-2 Performance Panel, SeraCare) were pooled in equal volumes and included on every plate as a control. Background values from no-serum wells were subtracted from the sample read-outs to give a net median fluorescence intensity (MFI) value. The net MFI value for each test sample was adjusted by dilution factor, such that antibody levels are represented as adjusted “units.” Baseline cut-offs for each antigen and antibody type were calculated using Receiver Operating Curve (ROC) analyses. The panel of 113 negative pre-pandemic sera were measured along with acute PCR-confirmed COVID-19 patients with sera collected 7-40 days post-infection (n=18), and to maximise sensitivity, the resulting 98% specificity cut-off for each antibody was taken as the baseline.

### Surrogate neutralisation assay

Surrogate neutralisation assays were performed using the SARS-CoV-2 sVNT (GenScript) according to the manufacturers’ instructions as previously described.^12,22^ Briefly, samples were diluted 1:20 and incubated with an equal volume of peroxidase-conjugated RBD for 30 min at 37°C, then added to wells pre-coated with human ACE-2 receptor protein and incubated for 15 min at 37°C prior to washing and development. Inhibition was calculated as (1—OD sample/OD of negative control) × 100. Any sample with a percentage inhibition ≥20% was deemed positive for NAbs, with this manufacturers cut-off previously shown to result in 100% specificity in our laboratory.^22^

### Mitra samples

To simulate remote blood sampling from a fingerprick using volumetric absorptive microsampling, Mitra devices (Neoteryx) were placed onto the surface of whole blood collected from an EDTA tube and filled. The devices were left to dry at room temperature and stored for a maximum of 3 weeks or frozen at −20°C until use. To elute the dried blood, the tip of the Mitra device was placed in 400ul of PBS +1% BSA + 0.05% Tween 20 and incubated overnight at 4°C, with shaking (300rpm). Eluents were stored at 4°C for up to one week or at −80°C until use. The ‘serum equivalent’ dilution of eluents was calculated to be 1:40, based on the assumption that serum constitutes 50% of the whole blood volume. To compare antigen-specific IgG levels in eluents with levels measured in serum, eluents were diluted appropriately and measured alongside matched serum samples in the multiplex assay. Overall differences in raw values between eluents and sera were low for each antigen (average CV= 8.6%, 11.9% and 9.2% for N, RBD and Spike).

### Statistical analysis

Data were analysed using Prism 8 (GraphPad) or R (version 4.0.2) within R Studio (version 1.2.5042) using packages rstatix (v 0.6.0; Kassambara, 2020) and the tidyverse (v1.3.0; Wickham, 2019). Locally estimated scatterplot smoothing (LOESS) with a span of 0.95 were used to visualise trends in antibody levels over time. Kruskal-Wallis followed by pairwise Dunn tests were performed to calculate statistical significance. Bonferroni method was used to correct for multiple comparisons. A *P*-value of ≤ 0.05 was considered statistically significant. The following models were applied to NAb data.

1. Exponential decay in which an exponential decay curve was fitted to the full dataset

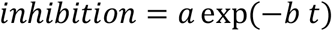

where *t* is measured in days. This method will likely underestimate the decay rate if the period in which antibodies increase after infection (growth period) for some individuals is long. It is also strongly affected by potential differences in both the size and timing of the peak level of NAbs between individuals. The best fit coefficients with 95% confidence interval are *a* = 66 (56.7, 75.3) and *b* = 0.0011 (0.00005, 0.00217).

2. Growth and decay, which uses a two-part model fitted to the full dataset

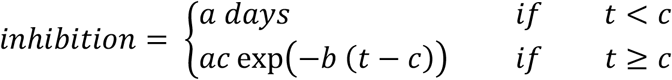

This allows an initial growth period to account for the underestimate that results from fitting a decay curve to measurements still in the growth phase. However, strong differences between individuals, particular in the timing of the peak level of NAbs will still affect estimations. This model predicts a slightly faster decay speed than the exponential decay model *b* = 0.0016 (0.00053, 0.00274) that starts after 11 days (*c* = 11 (5.65, 16.35)).

3. Individual decay model

Of the 189 data points for NAbs, 127 are multiple measurements from 50 individuals, of which 33 (88 measurements) are in the exponential decay stage, i.e. inhibition is decreasing with time. The individual decay model incorporates data from these 33 individuals only. An exponential decay curve is fitted to each individual separately. An exponential distribution is fitted to the individual decay rates and the best fit parameter and 95% confidence interval reported. The mean decay rate much is faster than predicted by the other models with *b* = 0.00476 (95% confidence interval 0.00348, 0.00691).

## Data Availability

The data that support the findings of this study are available on request from the corresponding author, NM, upon reasonable request.

## Acknowledgments and Funding

This work was funded by the School of Medicine Foundation (University of Auckland) and the COVID-19 Innovation Acceleration Fund (Ministry of Business, Innovation and Employment). We thank Ms Lauren Carlton and Dr Prachi Sharma for laboratory support and Linfa Wang at Duke-NUS and Genscript for providing sVNT testing kits and technical advice.

