## Supplementary information for "Comprehensive analysis of SARS-CoV-2 antibody dynamics in New Zealand"

**Supplementary Figure 1.** (A) Scatterplot showing an example correlation between Spike (S) protein IgG measured using the bead-based assay in single-plex and triplex with Nucleocapsid (N) protein and receptor binding domain (RBD), indicating compatibility of the antigen in a multiplex format. (B) Scatterplots showing the correlation between (i) N protein IgG measured in the triplex bead based assay and the Abbott Architect SARS-CoV-2 assay (n=141), (ii) RBD IgG measured in the triplex bead based assay and an in-house RBD ELISA (n=43) and (iii) S protein IgG measured in the triplex bead based assay and an in-house S protein ELISA (n=43), all using PCR-confirmed COVID-19 patients. The solid lines represent the linear regression equations. R squared and *P*-values are shown. MFI= median fluorescence intensity, AUC= area under curve.

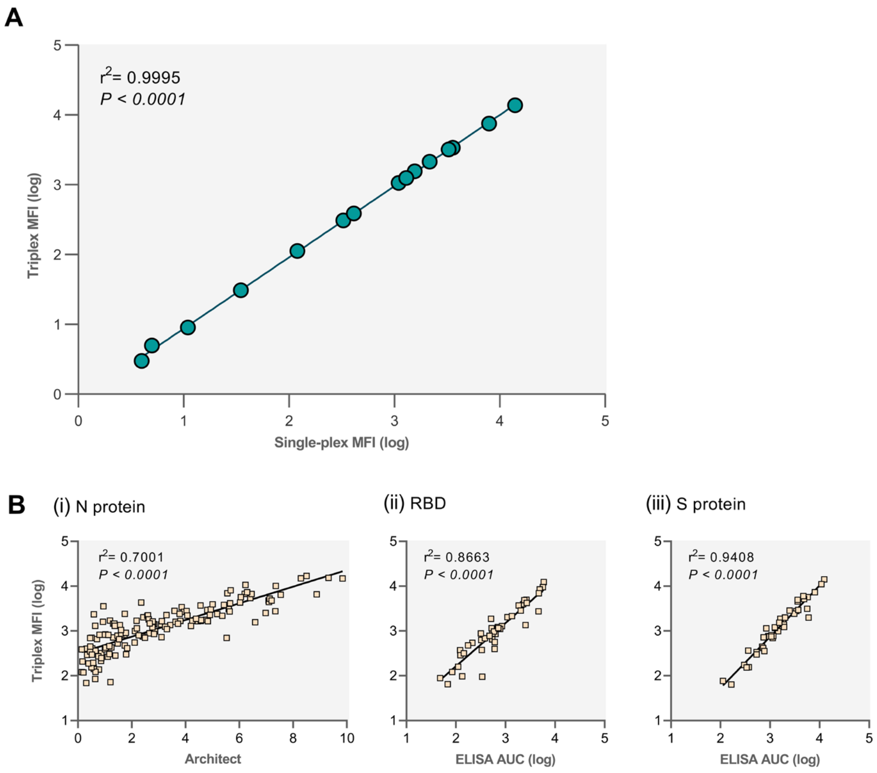

**Supplementary Table 1**. Percentage of PCR-confirmed COVID-19 samples that had antibody responses above calculated baseline levels. Isotype and subclass responses are shown for each antigen (N protein, RBD and S protein) across three time groups (7-62 days, n=27, 63-124 days, n=79, 125-250 days, n=80).

| | **IgG** | | | **IgA** | | | **IgM** | | | **IgG1** | | | **IgG2** | | | **IgG3** | | | **IgG4** | | |
|---|---|---|---|---|---|---|---|---|---|---|---|---|---|---|---|---|---|---|---|---|---|
| | 7-62 days | 63-124 days | 125-250 days | 7-62 days | 63-124 days | 125-250 days | 7-62 days | 63-124 days | 125-250 days | 7-62 days | 63-124 days | 125-250 days | 7-62 days | 63-124 days | 125-250 days | 7-62 days | 63-124 days | 125-250 days | 7-62 days | 63-124 days | 125-250 days |
| **N** | 85 | 82 | 54 | 48 | 9 | 1 | 11 | 1 | 0 | 70 | 53 | 30 | 52 | 28 | 18 | 67 | 52 | 23 | 48 | 24 | 10 |
| **% (n)** | (23/27) | (65/79) | (43/80) | (13/27) | (7/79) | (1/80) | (3/27) | (1/79) | (0/80) | (19/27) | (42/79) | (24/80) | (14/27) | (22/79) | (14/80) | (18/27) | (41/79) | (18/80) | (13/27) | (19/79) | (8/80) |
| **RBD** | 100 | 97 | 99 | 96 | 65 | 63 | 70 | 32 | 23 | 89 | 67 | 64 | 67 | 19 | 23 | 96 | 54 | 23 | 44 | 1 | 0 |
| **% (n)** | (27/27) | (77/79) | (79/80) | (26/27) | (51/79) | (50/80) | (19/27) | (25/79) | (18/10) | (24/27) | (53/79) | (51/80) | (18/27) | (15/79) | (18/80) | (26/27) | (43/79) | (18/80) | (12/27) | (1/79) | (0/80) |
| **S** | 100 | 96 | 96 | 81 | 44 | 43 | 67 | 27 | 14 | 85 | 67 | 59 | 67 | 25 | 21 | 100 | 84 | 60 | 52 | 11 | 3 |
| **% (n)** | (27/27) | (76/79) | (77/80) | (22/27) | (35/79) | (34/80) | (18/27) | (21/79) | (11/80) | (23/27) | (53/79) | (47/80) | (18/27) | (20/79) | (17/80) | (27/27) | (66/79) | (48/80) | (14/27) | (9/79) | (2/80) |

**Supplementary Figure 2**. Violin boxplot showing temporal neutralising antibody analysis. Boxes represent the 25th, median and 75th percentiles. Paired samples were available from a subset of participants over two or three time points (TP). Kruskal-Wallis followed by pairwise comparison (pwc) using Dunn test and Bonferroni correction for multiple comparisons indicated significant decrease of neutralising antibodies from TP1 to TP3, * = $P$ <0.05.

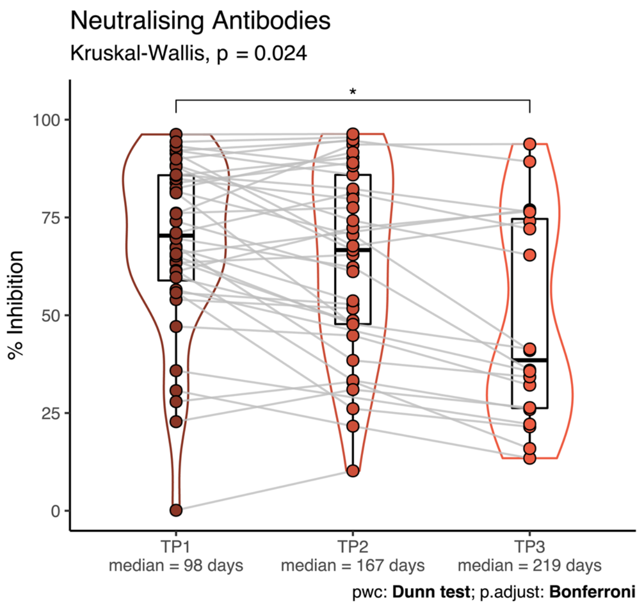

**Supplementary Table 2.** Summary of neutralising antibody (NAb) decay rate and half-life predicted from three different models

| | **Decay rate** (95% Confdence Interval) | **Expected half-life** (95% Confidence Interval) |
|---|---|---|
| Exponential decay | 0.00111 (0.00005 - 0.00217) | 625 days (319 - 13465) |
| Growth then exponential decay | 0.00163 (0.00053 - 0.00274) | 425 days (253 - 1316) |
| Individual decay | 0.00476 (0.00348- 0.00691) | 146 days (100 - 199) |
